# The Heart Health Yarning Tool: co-designing a shared decision-making tool for cardiovascular disease prevention and risk management

**DOI:** 10.1101/2024.11.19.24317586

**Authors:** Shannon McKinn, Judith Parnham, David Follent, Marguerite Tracy, Rosemary Wyber, Natasha Freeman, Rajesh Puranik, Michelle Dickson, Carissa Bonner

## Abstract

Due to the ongoing impact of colonisation, Aboriginal and Torres Strait Islander people live with a greater burden of cardiovascular disease (CVD) than non-Indigenous Australians. Shared decision-making is recognised as an essential component of person-centred care. However, the previous guidelines for CVD risk assessment and prevention did not engage Aboriginal and Torres Strait Islander people. Additionally, there has been a lack of tools to support clinician communication and shared decision-making to address CVD prevention in this important “at-risk” population. We developed the Heart Health Yarning Tool, an online shared decision-making resource co-designed with Aboriginal and Torres Strait Islander people, to be implemented alongside the new Australian guidelines for primary CVD risk assessment and management. This was a three phase project: Phase 1 consisted of a stakeholder consultation and co-design process, including consumer yarning workshops (n=21), individual yarning sessions with Aboriginal and Torres Strait Islander Health Workers/Practitioners (n=8), consumers (n=17), and interviews with general practitioners (n=5). Phase 2 involved a mapping process, where qualitative interview data was integrated into the conceptual framework of an existing culturally adapted shared decision-making model, ‘Finding Your Way,’ in order to tailor the model to the CVD context. Phase 3 involved developing and testing content for the new tool, based on findings from Phases 1 and 2, using evidence-based shared decision-making formats. The resulting tool supports health professionals to make shared decisions about heart health with Aboriginal and Torres Strait Islander people. It can be used as a conversation guide in primary care consultations or as a training tool for health professionals. Future research will assess whether use of the Heart Health Yarning Tool improves health professionals’ cultural and shared decision-making competencies as well as cardiovascular outcomes in Aboriginal and Torres Strait Islander people.

**KEY MESSAGES:** *What is already known on this topic:*

- Despite the fact that shared decision-making is recognised as an essential component of person-centred care, there are few culturally adapted shared decision-making resources
- Culturally specific resources are needed to support shared decision-making around heart health with Aboriginal and Torres Strait Islander people, a priority group for cardiovascular disease prevention

*What this study adds:*

- We developed an online shared decision-making tool to be used by primary care providers with the new Australian guidelines for primary cardiovascular disease prevention
- The Heart Health Yarning Tool was co-designed with Aboriginal and Torres Strait Islander people, incorporating culturally relevant content with evidence-based shared-decision making formats.

*How this study might affect research, practice or policy:*

- The Heart Health Yarning Tool can be used as a conversation guide, health consumer
- education resource, and as a training tool for health professionals.
- The Heart Health Yarning Tool may support health professionals to offer culturally responsive health care to Aboriginal and Torres Strait Islander people

## INTRODUCTION

The ongoing effects of colonisation result in Aboriginal and Torres Strait Islander Australians living with a high burden of cardiovascular disease (CVD). The prevalence of CVD is 1.3 times higher, and CVD mortality is 3 times higher for Aboriginal and Torres Strait Islander people than non-Indigenous people in Australia [1]. On average, Aboriginal and Torres Strait Islander people develop and die from CVD 10-20 years earlier than non-Indigenous Australians [2,3]. This gap is driven by the effects of colonisation, within and outside of the health system, and lack of access to culturally safe preventive health services [4–7] (see Box 1 for the Aboriginal and Torres Strait Islander primary health care context in Australia).

### Box 1

The Aboriginal and Torres Strait Islander primary health care context in Australia

#### Where do Aboriginal and Torres Strait Islander people access primary health care services?

Primary health care services for Aboriginal and Torres Strait Islander people are delivered by both mainstream general practice clinics, and Aboriginal and Torres Strait Islander-specific primary care organisations. As of 2023, there were 213 organisations providing Aboriginal and Torres Strait Islander-focussed primary health care, of which 69% were Aboriginal Community Controlled Health Organisations (ACCHOs) [8]. In 2022-2023, these organisations served 443,000 Aboriginal and Torres Strait Islander clients [8], representing 45% of the total population of Aboriginal and Torres Strait Islander people [9].

#### Who are Aboriginal and Torres Strait Islander Health Workers/Practitioners?

Aboriginal and Torres Strait Islander Health Workers/Practitioners are one of the only ‘identified’ roles within the health care system in Australia, meaning they must be filled by an Aboriginal and/or Torres Strait Islander person. They are typically located within primary care services [10]. Both Health Workers and Health Practitioners are required to hold specific Aboriginal and Torres Strait Islander healthcare qualifications. Aboriginal and Torres Strait Islander Health Practitioner is a protected title and nationally registered health profession under the Aboriginal and Torres Strait Islander Health Practice Board [11].

Aboriginal and Torres Strait Islander Health Workers/Practitioners provide a wide range of clinical and non-clinical services to support the delivery of holistic and culturally safe primary health care within their communities [12]. Aboriginal and Torres Strait Islander Health Workers/Practitioners are one of the few roles within the health care system with an explicit responsibility to advocate for culturally safe care at the whole-of-service level [10,13].

Primary prevention is the mainstay of preventing CVD for individuals. Comprehensive risk assessment allows for calculation of CVD risk based on composite risk factors and offers risk-stratified management approaches. In Australia updated CVD prevention guidelines were released in July 2023 providing new guidance about CVD risk assessment [14]. These guidelines include specific guidance for healthcare professionals working with Aboriginal and Torres Strait Islander Australians, including assessing individual risk factors at an earlier age and potentially reclassifying to higher risk category [2]. This further develops recommendations made in the previous national CVD prevention guidelines from 2012 [15]. However, there remain substantive challenges in offering and operationalising CVD risk assessment for Aboriginal and Torres Strait Islander people. To date there is a paucity of specific CVD screening and risk stratification tools designed specifically to meet the needs of Aboriginal and Torres Strait Islander Australians [3].

Shared decision-making (SDM) is increasingly recognised by clinicians and policy makers as an essential component of person-centred care [16,17]. SDM involves both the clinician and the patient being involved in making key decisions about the patient’s health care plan and treatment, based on their personal preferences and values. SDM involves the patient and clinician working in collaboration in order to enable a patient-centred style of communication and care [17]. There is evidence that SDM is associated with improved patient health, knowledge and satisfaction outcomes occurring as a result of reduced decisional conflict and clinician controlled decision-making, and enhancing patient risk perception accuracy [18–20]. Despite this, evidence shows that clinicians are less likely to engage in SDM with people from culturally and linguistically diverse backgrounds [21].

Few culturally adapted SDM models exist [22,23]. However, during the COVID-19 pandemic, the New South Wales Department of Health (NSW Health)) partnered with Aboriginal communities to develop a new SDM model [24,25]. ‘Finding Your Way’ is a culturally adapted SDM model that was co-designed with Aboriginal Health Workers/Practitioners (see Box 1) and community members to respond to the need for SDM tools in the context of COVID-19 vaccination [24–26]. It is the first and, to our knowledge, only culturally adapted SDM resource for Aboriginal people in Australia [25]. The ‘Finding Your Way’ model (See Figure 1) consists of eight inner circles that represent the core elements of SDM, surrounded by interconnected concepts that represent the scaffolding to support Aboriginal and Torres Strait Islander people as they make health decisions. At the centre of the model is ‘Physical, Social, and Emotional Wellbeing,’ representing a holistic conceptualization of Aboriginal and Torres Strait Islander health and wellbeing [26]. While developed in the context of COVID-19, acceptability and usability testing of the original model suggested that it can also be adapted for use for other health and wellbeing contexts [25].

**Figure 1:**
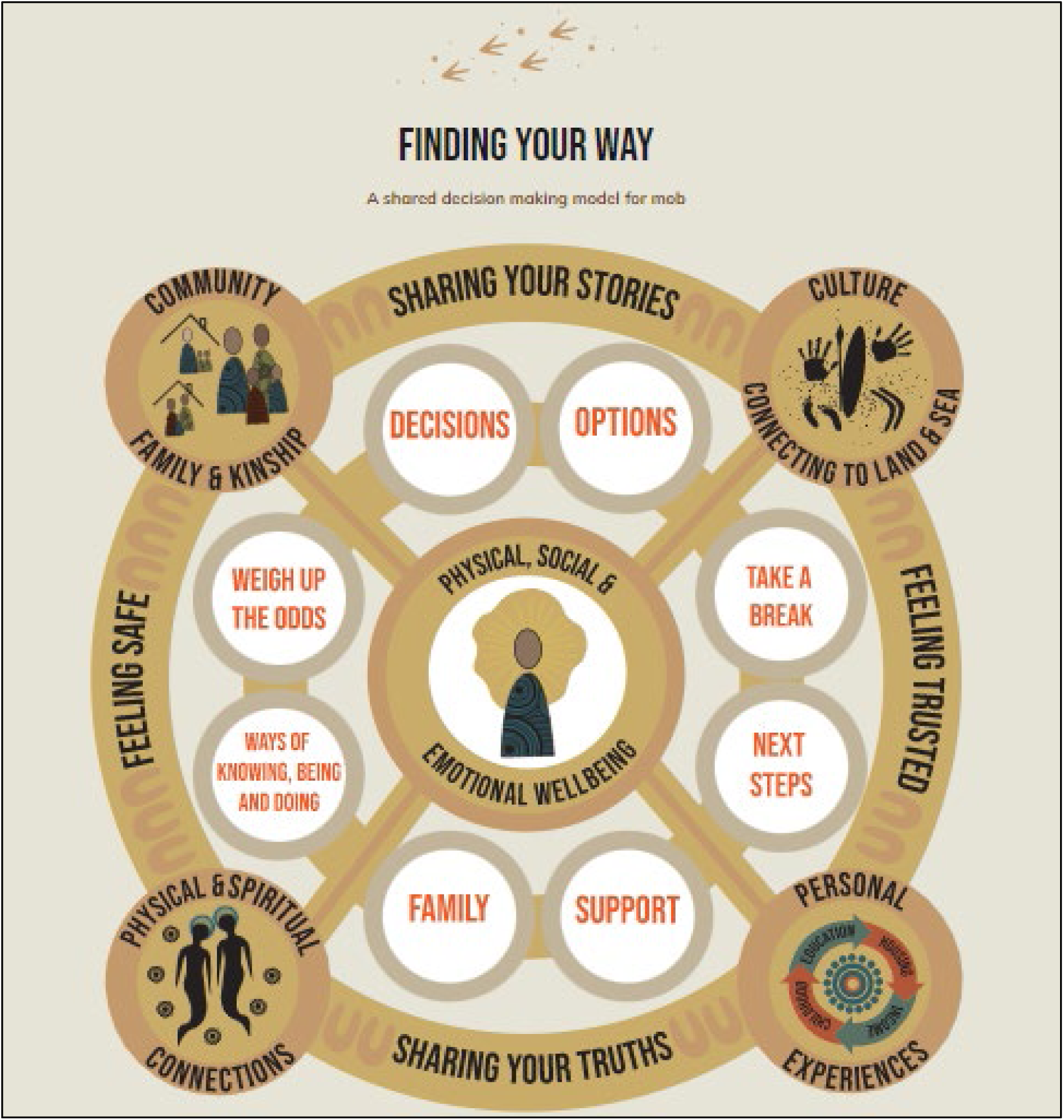
‘Finding Your Way’ – A Shared Decision Making Model for Mob [25,26]. Artwork by Belinda Coe

The previous 2012 guidelines for CVD risk assessment and prevention did not engage Aboriginal and Torres Strait Islander people. Aboriginal and Torres Strait Islander Health Workers/Practitioners had no active role in development of the guidelines [27] nor in the parallel development of GP and consumer SDM tools [28], leaving a lack of tools to support clinician communication and SDM with this priority “at-risk” population for CVD prevention [29]. Our previous research has identified that diverse and culturally specific resources and implementation pathways may be needed to support SDM with Aboriginal and Torres Strait Islander communities. We have identified clear gaps that potentially affect access to preventive health services for CVD, including leaving Aboriginal and Torres Strait Islander Health Workers/Practitioners out of Heart Health Check models of care and identifying a lack confidence and competence to deliver culturally responsive care in mainstream general practice staff [27,30].

Co-design involves collaboration between stakeholders, including service users, researchers, and implementers to develop, implement and evaluate solutions to real-world problems, with a focus on consumers’ insights [31,32], and their expertise based on their own lived experience [33]. This paper describes the co-design and development process of the Heart Health Yarning Tool, a SDM tool/conversation guide for Aboriginal and Torres Strait Islander people, adapted from the ‘Finding Your Way’ model. It is to be implemented alongside the new Australian guidelines for primary CVD risk assessment and management.

## METHODS

### Study design

This was a multi-phase project encompassing three phases (Figure 2): consultation, mapping, and development. Phase 1 consisted of a stakeholder consultation and co-design process, centred on the use of a yarning approach with Aboriginal and Torres Strait Islander participants, and semi-structured interviews. Yarning is a method of having conversations and sharing stories in a culturally supported manner that prioritises Aboriginal and Torres Strait Islander ways of communication [34,35]. The yarning approach used in this study has been described elsewhere [27]. We conducted consumer yarning workshops, and individual yarning sessions with Aboriginal and Torres Strait Islander Health Workers/Practitioners and consumers, in-depth interviews with general practitioners (GPs), and user testing interviews with Aboriginal and Torres Strait Islander consumers. Phase 2 involved a mapping process, where yarning and interview data was mapped onto the conceptual framework of the ‘Finding Your Way’ model, in order to tailor the SDM model to the CVD context. Phase 3 involved developing and testing content for the new SDM tool, based on findings from Phases 1 and 2, using evidence based SDM formats.

**Figure 2.**
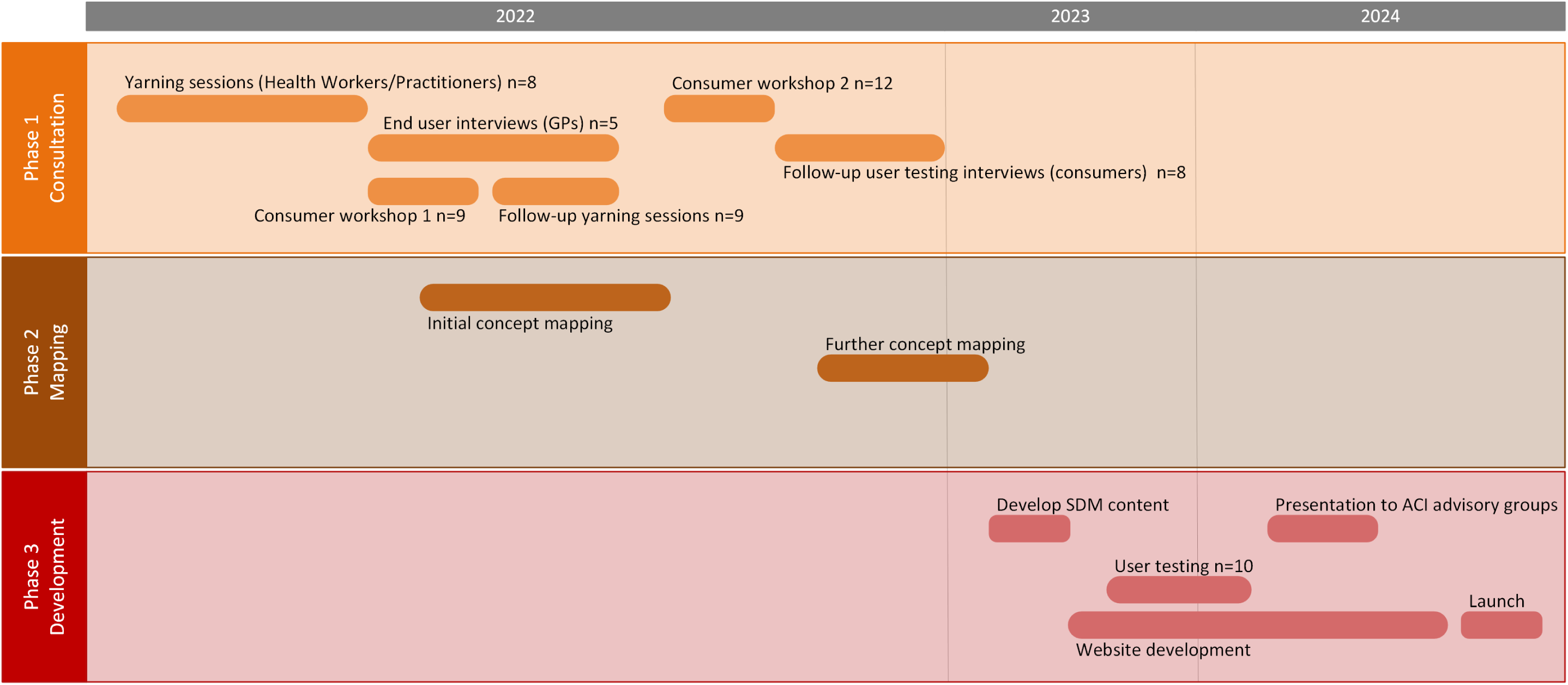
– Phases of co-design and Heart Health Yarning Tool development process. GP: general practitioner, ACI: Agency for Clinical Innovation, SDM: shared decision-making

### Author positionality

The study was designed and conducted by a team including Aboriginal (DF, MD), Aboriginal and Torres Strait Islander (JP), and non-Indigenous researchers (SM, RW, NF, RP, MT, CB).

#### Aboriginal and Torres Strait Islander authors

JP is an Aboriginal and Torres Strait Islander Health Worker. She is a an Ankamuthi and Erub descendant. DF is Senior Project Officer in Chronic Care for Aboriginal People at the Agency for Clinical Innovation, NSW Health. He is a proud Bundjalung man. MD is an associate professor and Director of the Poche Centre for Indigenous Health at the University of Sydney. She is a Darkinjung/Ngarigo Aboriginal Australian.

#### Non-Indigenous authors

SM is a research fellow. MT is a senior lecturer and GP. RW is a research fellow and GP. NF is a GP. RP is a professor and consultant cardiologist. CB is an associate professor and behavioural scientist.

### Setting

During February – November 2022, we conducted Phase 1 stakeholder consultation with participants across five Australian states (New South Wales, Queensland, South Australia, Victoria, Western Australia). Phase 2 mapping occurred throughout 2022 and 2023, with Phase 3 development and user testing through 2023 to May 2024.

### Participants and recruitment

Participants in the co-design process included Aboriginal and Torres Strait Islander consumers, Aboriginal and Torres Strait Islander Health Workers/Practitioners, and GPs. Aboriginal and Torres Strait Islander Health Workers/Practitioners were recruited through the National Australian Association of Aboriginal and Torres Strait Islander Health Workers and Health Practitioners. Consumers were recruited for yarning workshops via a referral process from Aboriginal and Torres Strait Islander Health Worker/Practitioners and the networks of the research team. Consumers who participated in a workshop were subsequently invited to participate in an individual interview. General practitioners were recruited via the networks of the research team.

All participants received online gift vouchers for their attendance at workshops (250 AUD) and individual interviews (75 AUD). This amount aligned with recommendations from Health Consumers NSW [36].

### Data collection

Phase 1 consisted of eight yarning sessions with Aboriginal and Torres Strait Islander Health Workers/Practitioners, two group yarning workshops with 17 individual follow up sessions with Aboriginal and Torres Strait Islander consumers, and five semi-structured interviews with GPs.

Yarning sessions with Aboriginal and Torres Strait Island Health Workers/Practitioners were conducted by an Aboriginal and Torres Strait Islander member of the research team (JP) who is also an Aboriginal and Torres Strait Islander Health Worker. These sessions covered awareness of current CVD screening tools, the role of the Aboriginal and Torres Strait Islander Health Worker/Practitioner in CVD screening, and participants’ opinions of the current screening tool [28] and its efficacy for clients and clinicians.

Consumer yarning workshops were held online, via Zoom, in order to facilitate participation by consumers across varied geographical locations. The workshops were led by three Aboriginal and Torres Strait Islander members of the research team (JP, DF, MD). The yarning workshops included a group discussion about making decisions around healthcare, and (voluntary) sharing of personal experiences of SDM and patient-centred care. They also included demonstrations and discussions of two SDM resources, (i) a patient risk assessment tool our team developed for the 2012 Australian CVD risk management guidelines aimed at the general population [29], and (ii) the culturally adapted ‘Finding Your Way’ SDM model [25,26]. Finally, consumers discussed what tools they thought would be most useful for them to support the new CVD prevention guidelines. Following each of the two yarning workshops, the workshop participants were invited to participate individually. After the first workshop, JP conducted a round of individual consumer yarning sessions, focusing on personal experiences of SDM and person-centred care. A second round of consumer interviews were conducted by a non-Indigenous researcher with expertise in qualitative research and SDM (SM), focused on eliciting feedback on existing resources using a think aloud method [37]. Participants were encouraged to say their thoughts out loud as they interacted with the ‘Finding Your Way’ website, and prompted to reflect on the relevance of specific SDM elements to CVD prevention and management, in order to gain concurrent insight into their thoughts and impressions of the model [37,38].

Non-Indigenous researchers conducted the GP interviews (SM, NF). GP interviews covered GPs’ use of SDM and decision support tools, patient responsiveness to SDM, experiences of SDM with Aboriginal and Torres Strait Islander patients, and barriers and facilitators to using SDM approaches in practice. A second round of GP interviews was conducted during Phase 3 to record user feedback on the prototype of the Heart Health Yarning Tool, using the think aloud method described above.

All yarning and interview sessions were conducted online via Zoom video or voice call, and were audio-recorded and transcribed, with participants’ written consent. The topic guides used for yarning and interviews are available in Supplementary file 1.

### Data analysis

The initial yarning sessions were analysed using a verbal yarning approach to develop themes around SDM (JP, MD, CB; SDM themes reported separately [27]). We then adopted a Framework Analysis approach [39] to map all data to the a priori categories of the ‘Finding Your Way’ model. Data from all three participant groups was coded and categorised in NVivo [40], and then mapped in a framework, with participants as rows and ‘Finding Your Way’ concepts as columns. The data for each concept was summarised for discussion with all members of the research team as part of the process of developing CVD specific content for the Heart Health Yarning Tool.

### Public and patient involvement

*Thiitu Tharrmay* Aboriginal and Torres Strait Islander Reference Group, is convened by Yardhura Walani at Australian National University. *Thiitu Tharrmay* members are Aboriginal and Torres Strait Islander peoples who are consumers or providers of healthcare, with knowledge and/or experience with research and health policy [41]. *Thiitu Tharrmay* advised on appropriate conduct of this research, and provided comment on the research methods, including recruitment and data collection methods. Aboriginal and Torres Strait Islander researchers were involved in all stages of the project, including study design, data collection, analysis, interpretation, writing and revising the manuscript (JP, DF, MD). Aboriginal and Torres Strait Islander consumers were involved in the co-design and user testing of the Heart Yarning Tool.

### Ethics statement

Ethics approval was obtained from the Australian Institute of Aboriginal and Torres Strait Islander Studies (EO294-20210826) after input from *Thiitu Tharrmay*. All participants provided signed consent to participate.

## RESULTS

### Participant characteristics

To ensure participant privacy, and in accordance with advice from *Thiitu Tharrmay* Aboriginal and Torres Strait Islander Reference Group, individual demographic details of participants were not formally collected. This decision reduced potential for participants to be identified, particularly within a relatively small professional pool of Aboriginal and Torres Strait Islander Health Workers/Practitioners. A consistent approach was used for all participant groups. Participants’ age ranged from in their 20s to 60s, and there was a mix of gender diversity, living/working in metropolitan and rural locations.

### Phase 1: Stakeholder consultation and co-design

Most of the participating Aboriginal and Torres Strait Islander Health Workers/Practitioners found the CVD risk calculator [15] easy to use and helpful in developing further conversations with their clients about CVD prevention. However, participants also noted that important economic, cultural and environmental factors for Aboriginal and Torres Strait Islander people were not considered. Participants also raised concern about the health literacy demands of interpreting the results of the calculator.

> *“I think it’s a good tool because it’s very easy to use and the results you get back quick, but it doesn’t also accommodate for our mob that Aboriginal and Torres Strait Islander people have shorter life spans than non-Indigenous people, and are facing different health factors and social and economic hardships or barriers that also come into play for our health. (…) I do not feel that it’s appropriate. It’s not targeted for Aboriginal and Torres Strait Islander people. A more targeted tool would be more appropriate.”* (Aboriginal and/or Torres Strait Islander Health Worker/Practitioner)

Aboriginal and Torres Strait Islander consumers gave positive feedback on the format of the yarning sessions, stating that they appreciated the space to share stories about health journeys and health care experiences with each other.

> *“Sometimes you think you know like you’re the only one that’s going through something but there’s other people who can relate and connect to that experience and (…) share those ideas around that shared decision-making and how we can be in control of our own health journey (…) I thought that was a really good eye-opener.”* (Consumer reaction to Yarning workshop)

The workshops were used to both present and discuss the patient risk assessment tool [29] and ‘Finding Your Way’ [25,26] models. Consumer participants expressed concerns about the complexity of current CVD tools; this was a particular issue for members of the community with limited health and digital literacy. Echoing concerns already raised by Aboriginal and Torres Strait Islander Health Workers/Practitioners, several consumers mentioned factors that they believe strongly impact CVD risk in their communities but are not accounted for in current biomedical risk models. These included socioeconomic circumstances, poor mental health, and racism. Consumers liked the appearance of the ‘Finding Your Way’ model, particularly liking the non-linear and interconnected nature of the visual layout, and the focus on a holistic concept of health and wellness that encompassed physical, social and emotional wellbeing. Consumers also reacted well to certain visual elements of the patient risk assessment tool, specifically the icon array.

> *“Aesthetically, the circles are really good. We tend to think in the sense – well, where I’m from anyway – circles are representative of processes, we don’t run linear to everything, everything is connected somehow, sometime, so this is a really good representation. And the connection to many of the themes, like community, family, and culture, it’s understandable, I can understand it, I can see how it’s all connected just by virtue of the conceptual arrangement.”* (Consumer reaction to ‘Finding Your Way’ model)

> *“I liked the visual, I liked the little men, the little people in the little chart - I loved that. It really got to me. With the 100 people I really liked that.”* (Consumer reaction to patient risk assessment tool)

General practitioners emphasised the importance of using a SDM approach to CVD risk management with Aboriginal and Torres Strait Islander people, both from a human rights perspective and also in terms of achieving a successful outcome. GPs voiced similar concerns to the other participant groups about factors that go unaccounted for in current CVD risk calculators.

> *“You’re more likely to get an actual result if you’ve made decisions jointly. There’s no point in just deciding on behalf of a patient what they need to do when they definitely have no intention of doing it and also when you haven’t explored the reasons for why they may choose to make health decisions or not. But I mean it’s also just a human rights issue. We all have the right to be in charge of our own health, I think.”* (General practitioner)

> *“It’s a bit sort of general, or blunt in a way, in the way it stratifies a patient, because it doesn’t take into account things like family history, or alcohol status, presence of chronic kidney disease, and I think those – and even just the patient’s diet and lifestyle.” (General practitioner)*

### Phase 2: Concept mapping

During Phase 2 we conducted two rounds of a deductive mapping process to map interview data to the concepts of the ‘Finding Your Way’ model. We conducted an initial mapping exercise after the first round of consumer yarning interviews was completed. This exercise incorporated data from all three participant groups: Aboriginal and Torres Strait Islander Health Workers/Practitioners, GPs and consumers. The first round of mapping revealed gaps in our data around specific concepts in the ‘Finding Your Way’ Model. The second round of consumer user testing interviews responded to these gaps by directing the discussions to focus on specific concepts while participants used the ‘Finding Your Way’ tool. Particular focus was placed on the inner circle of eight SDM concepts in order to tailor content around CVD risk prevention and management. Examples of interview data mapped to the ‘Finding Your Way’ concepts are shown in Table 1.

**Table 1:** Examples of interview data mapped to Finding Your Way (FYW) shared decision-making processes.

| **FYW step/process** | **FYW description** | **Example of mapped data / Illustrative quote** | **Heart Health Yarning Tool content** |
| --- | --- | --- | --- |
| Options | Yarn about the options and the benefits and risks of treatment. Ask questions, share knowledge and feelings about the treatment options | *The options gives us control, it gives us that power over our own – making decisions about our own treatment and what we want and our own needs, and I really like that.* | Which options do you want to consider to improve your heart health? |
| Support | Yarn about what is happening today, including social, emotional and wellbeing needs and supports | *We used to go to walking groups, and they said you might be interested in endocrinologist via telehealth, and it's been beneficial. It's something that I would not have considered, and I found it a very beneficial referral.* | You have more in your life than just your heart health needs, talk about what else is important to your social and emotional wellbeing.<br>You can yarn to family, friends, health professionals and other people in your community about what you might need. Support can come from a team of health professionals, not just your health worker or doctor.<br>It's important for the people who are looking after you to know what works well for you, to support you in the best way for you. |
| Family or friends | Yarn about family and mob. If needed, bring others into the yarn for support. | *If you're going to give positive feedback to family about your experience, they're going to feel more entrusted to go as well and see about their heart. Yeah, so I think it's a really good tool and I think it just outlines the importance of yarning.* | Talking about your family and friends, your country and their experiences of heart and vascular issues can be helpful.<br>Yarn with your family and friends about your options. It is okay to make these decisions together with family. |
|  |  |  | Making decision with others is ok. You can bring family or friends to the doctor to help you make decisions.<br>You might all make some changes together to improve your health e.g. you might all go for a walk together every day.<br>Your family may have had to find solutions to improve their health in the past which may work for you too. They may have some tips they can share with you. |
| Ways of knowing, being and doing | Yarn about ways of knowing, being and doing to inform health decisions based on a person's values and beliefs | *When I went over to one of our clinics, I really felt heard, I really felt respected, it wasn't rushed and they actually sat down and treated you like a human being. They didn't lecture you. They actually talked about looking at our strengths as to how we can improve our health and focusing on what was wrong type thing. So, you felt like a human being.* | You are the expert about your story and your body. Nobody knows your journey better than you do, and everyone's journey is different. Here are some questions you can yarn about with your health professional to help them understand what's important to you. |
| Weigh up the odds | Yarn about the possible benefits and risks. Compare options and weigh up the odds individuals and for family and mob | *You can ask questions. What are my options? What are the benefits and harms? How likely are they to happen to me?* | There are different ways that you can improve your heart health. There will be good and bad things about each of these options. Think about which heart health options are best for you, your personal circumstances and what support you have from family, friends and your community. |
| Take a break | Yarn about taking a break (if needed) and making the decision later. Come back and yarn on another day | *You're not going to get all this done in one session. So, being able to have that follow-up, being... having that ongoing care, having the ongoing talking about, sharing our stories – having something you can take away and being followed up is very important.* | You might need more time to think about the options, or yarn with family and friends. It's ok to come back another time if you're not ready to make a decision yet. If you don't feel like you're in a safe place to yarn with someone you trust, you can talk to a different health professional. You can ask about seeing an Aboriginal and Torres Strait Islander Health Worker or Practitioner if you don't feel comfortable with your GP or nurse. You can ask to see a male or female health professional to help you feel more comfortable. |
| Next steps | Yarn about the next steps, including how and what to do next and what might get in the way. Follow up later | *This is what stuck with me, that he was naturally curious, and he actually asked me "What are the challenges, to you, getting a screen right here, right now?" Instead of giving me a lecture what he was saying was, "What can I do to help you to consider a screen?"* | When you are trying to make changes for your health some things work well and some things take more time to put into place. Some things you try may not work at all for you, at first – talk to your healthcare team and family about how they can support you to help you find what is best for you. Checking in is important and bring the people who support you on your health journey. |
| Decisions | Yarn to bring it all together and either decide to act now if ready, or wait | *I'd like to hear from a doctor's point of view obviously because that's their profession, but I also want them to hear what I'm saying and be* | Think about the lifestyle or medication options you want to consider. Yarn with your health |
|  |  | *able to answer it in a civilian way so it's understandable... not just chucking big words around... make you understand so you don't go home with a worried mind.* | professional about which ones might be best for you right now. |

### Phase 3: Heart Health Yarning Tool content and website development

At the start of Phase 3, a co-design workshop was held to draft content for the Heart Health Yarning Tool. Content was based on yarning/interview findings, concept mapping results, and key participant quotes. Participants in the workshop included three Aboriginal and Torres Strait Islander community members (one with lived experience of a heart condition, one Aboriginal and Torres Strait Islander Health Worker, one Indigenous health promotion academic), three non-Indigenous health professionals with experience in Aboriginal and Torres Strait Islander health (three GPs, one cardiologist), and two SDM researchers.

The CVD specific content was then implemented within evidence-based SDM formats, under the “weighing up the odds” component of ‘Finding Your Way.’ These formats included (i) a question prompt list [42] based on ‘Finding Your Way’ principles [25]; (ii) action planning tools to encourage adoption of habit changes around smoking, exercise and diet which were shown to enable lifestyle change in a previous trial [29]; and (iii) decision aids for blood pressure and cholesterol lowering medications which were shown to improve both GP and patient understanding of CVD risk [28,29] (see Figure 3 for overview, or Supplementary file 2 for full content).

**Figure 3:**
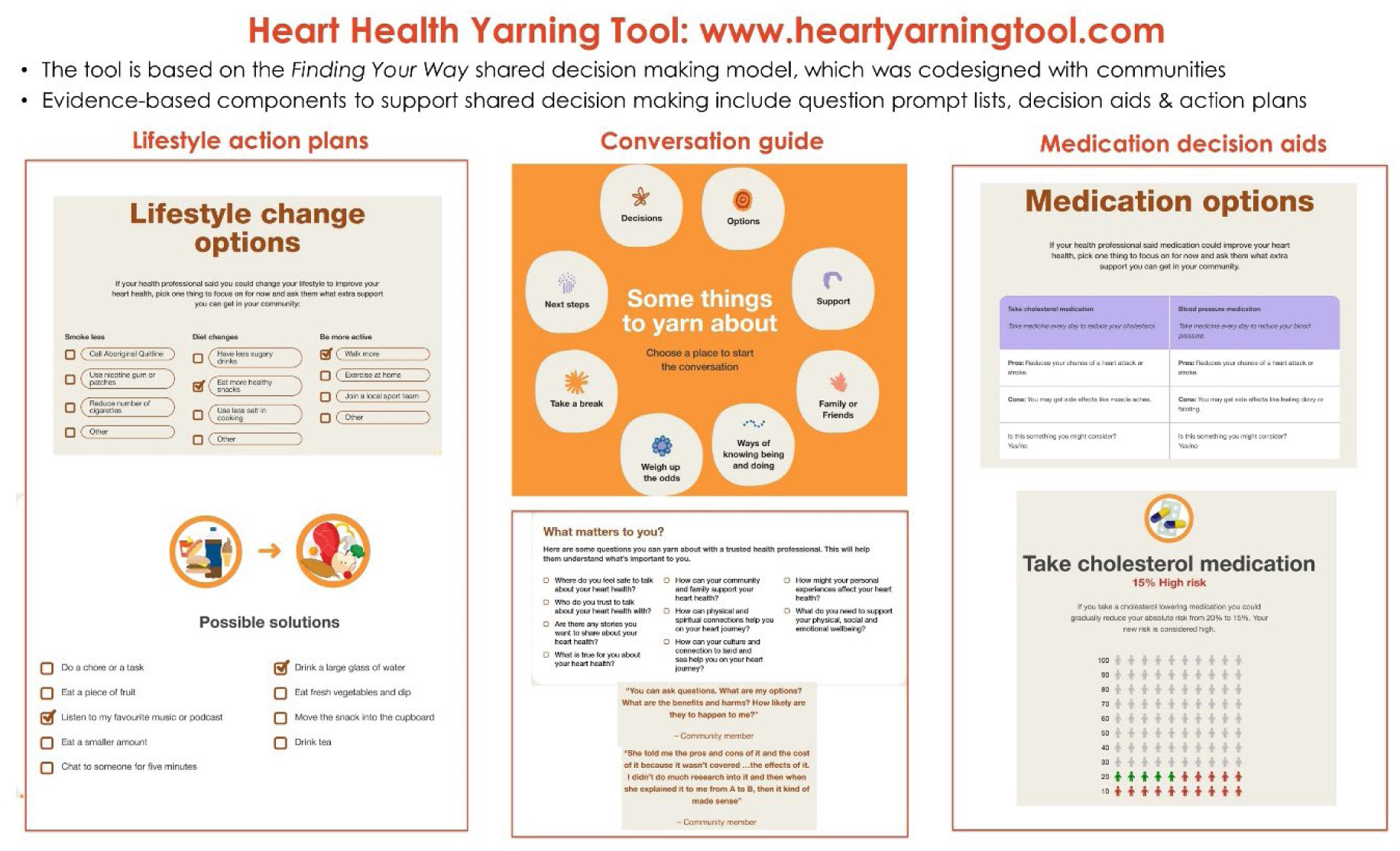
Heart Health Yarning Tool structure and components.

The website was developed by Aboriginal and Torres Strait Islander design company, Saltwater People, using artwork designed specifically for the broader CVD risk project. This was in response to feedback from community members requesting culturally appropriate visual designs across the interactive lifestyle and decision-making components of the tool.

User testing of the website was conducted by Saltwater People with 10 participants, including Aboriginal and Torres Strait Islander consumers and health professionals involved in the original yarning and interview sessions. Additional feedback was obtained from two advisory groups to the NSW Agency for Clinical Innovation: the Cardiac Network and the GP advisory group. Changes were made to the web tool in response to user feedback, including:

- wording clarification throughout the website
- a shortened two-page patient summary PDF about CVD risk
- more context for the use of the tool by health professionals on the home page
- a short demonstration video explaining key elements
- downloadable PDF summaries for every component for community members to take home from the consultation.

## DISCUSSION

This paper outlines the co-design process used to develop the Heart Health Yarning Tool (heartyarningtool.com) which can be utilised as a conversation guide in consultations, a patient education resource, and as a training tool for health professionals. The tool has been co-designed with end-users and Aboriginal and Torres Strait Islander community members. It was based on the ‘Finding Your Way’ model, which was also co-designed with Aboriginal people. In developing the tool, we have drawn on community and Aboriginal and Torres Strait Islander Health Worker/Practitioner perspectives to create content that may support primary care providers to offer culturally responsive care during discussions of cardiovascular risk. Ultimately, we hope that the tool will contribute to achieving improved heart health outcomes, that will subsequently contribute to addressing “the health inequity gap” that exists between Aboriginal and Torres Strait Islander and non-Indigenous people.

Interventions designed within community engagement models have been found to improve health outcomes and health behaviours in communities experiencing disadvantage and other challenges [43]. Globally, co-design has been successfully used to involve Indigenous communities in the research process, and to improve health services [31,33]. The biomedical principles of CVD prevention with Aboriginal and Torres Strait Islander people are not unique, and focus on healthy lifestyles, smoking cessation, and uptake of blood pressure and cholesterol-lowering medications. However, the factors that influence how and whether those actions can be put into practice are highly context-informed and culturally specific [31]. Qualitative research with Aboriginal women on their views of cardiovascular health and risk factors has emphasised the gap between biomedical and behavioural models of CVD prevention. The women’s focus on psycho-cultural-social factors supported the need for models of care that foster both physical and spiritual health, and ensure connection to family and community [44]. This highlights the critical importance of co-designed and culturally adapted tools such as the Heart Health Yarning Tool, that support health professionals to yarn with Aboriginal and Torres Strait Islander people about the sociocultural factors that affect their decisions about their heart health, which are not considered in mainstream CVD risk assessment and communication tools.

There is complex relationship between SDM and the delivery of culturally responsive health care. Cultural responsiveness and cultural safety are closely related, yet distinct, antiracism approaches to improving healthcare delivery and achieving equitable health outcomes [45]. While these terms are sometimes used interchangeably, cultural safety focuses on the subjective experience of the healthcare consumer [45]. Healthcare providers can strive to offer culturally responsive care, however only the end user of that care can determine whether a clinical interaction is safe [46,47]. Therefore, in the following paragraphs we use the term ‘culturally responsive’ when discussing the delivery of health care, and ‘culturally safe’ in reference to consumers’ lived experience of health care. In order to offer culturally responsive care, healthcare providers must make their own cultural background, and its impact on their clinical interactions, a focus for critical reflection [46,48]. Culturally responsive care acknowledges the barriers to effective care that arise from the inherent power imbalance between health care providers and health care consumers [49], seeks to redress this power imbalance through critical reflection [48], and focuses on *how* care is delivered, experienced and perceived, rather than *what* care is provided [47].

Previous research with Aboriginal and Torres Strait Islander people in Australia and Indigenous Peoples in Canada has demonstrated that cultural safety, along with trust and respect, are crucial prerequisites for SDM to occur [25,50,51]. Globally, Indigenous patients are more likely to engage in SDM and have greater trust in their health care provider when health information has been translated using culturally appropriate knowledge [22]. A systematic review of the literature on what constitutes culturally safe health care practice from the perspective of Aboriginal and Torres Strait Islander people found that across all health care settings studied, health systems were generally perceived to be disempowering and culturally unsafe for Aboriginal and Torres Strait Islander people [52]. A lack of partnership between provider and patient via SDM, as well as the absence of respect and trust, compromises the delivery of culturally responsive health care, and contributes to care being experienced as culturally unsafe [21,46].

### Strengths and limitations

Strengths of this work include the use of Indigenous research methods, specifically, the use of Aboriginal and Torres Strait Islander culturally appropriate processes for data collection, and leadership of Aboriginal and Torres Strait Islander members of the team in facilitating yarning sessions. Data collection was conducted online due to restrictions posed by the COVID-19 pandemic. This gave the project wider geographical reach, but also imposed limitations, in that workshop participants were not able to have “hands on” interaction with SDM tools, and the online method may have prevented rapport building, especially in group conversations. While the workshops and interviews comprised gender diversity, the majority of participants did identify as female. Following Aboriginal and Torres Strait Islander cultural protocols, separation of participants into Men’s and Women’s groups may facilitate more participation in future research, and elicit unique perspectives from each group that were not able to be elucidated in mixed gender groups [53].

## Conclusion

While the Heart Health Yarning Tool has been found to be acceptable to consumers and health professionals, additional training for health professionals is needed to support their use in practice, particularly for health professionals with less experience in Aboriginal and Torres Strait Islander health-care settings. Further research is needed to develop training support, and to evaluate SDM resources that are experienced as culturally safe by Aboriginal and Torres Strait Islander people and to identify appropriate implementation pathways.

## Data Availability

In line with our ethics approval, data from this study are not publicly available in order to protect the confidentiality of participants.

## Acronyms

CVD: Cardiovascular disease
FYW: Finding Your Way
GP: General practitioner
NSW: New South Wales
SDM: Shared decision-making

## Author contributions

SM, JP, MD, and CB conceptualised and designed the study. SM, JP and NF collected data. SM and JP analysed the data. SM drafted the manuscript. SM, JP, DF, MT, RW, RP, MD and CB contributed to data interpretation. All authors critically revised the manuscript and approved the final version for publication.

## Acknowledgements

We thank the members of *Thiitu Tharrmay* for their advice on the study design, and the community members and health professionals who participated in the study. We thank Saltwater People for their work designing and building the Heart Health Yarning Tool. We thank Carys Batcup for assisting with the ethics approval process, and Uday Yadav, Chelsea Liu and Deborah Wong for their assistance in delivering and disseminating the Heart Health Yarning Tool. We also thank Jason Agostino for foundational insights at the inception of this project.

