## Supplementary file 1 for "The Heart Health Yarning Tool: co-designing a shared decision-making tool for cardiovascular disease prevention and risk management"

Supplementary file 1: yarning and interview topic guides

### ABORIGINAL AND TORRES STRAIT ISLANDER HEALTH WORKER/PRACTITIONER TOPIC GUIDE

**Key topics (and prompts)**

- Awareness of current recommended CVD screening tools for Aboriginal and Torres Strait Islander Health Workers/Practitioners
    - How are clients CVD screened already in their community/facility.
    - Can you tell me how Aboriginal, and Torres Strait Islander Health Workers / Practitioners are currently utilised in the CVD screening process?
    - What is good about the current CVD screening tool and why?
    - Do you feel that the current CVD tool is culturally appropriate and why?

- Role of Aboriginal and Torres Strait Islander Health Worker/Practitioner in CVD screening
    - How does the role of the Aboriginal and Torres Strait Islander Health Worker /Practitioner operate for CVD screening in your workplace?
    - Do you find the current CVD screening tool allows you to do opportunistic screening in the community /primary health care setting and why?
    - Do you think that current CVD screening tool is at a level that is user friendly to most people and why?
    - Which MBS item numbers could possibly be claimed in the CVD screening process and by whom?

- Perceptions of current CVD screening tool and efficacy for clients and clinicians
    - What barriers have you found in the current CVD screening process?
    - What barriers have you found in the current CVD tool?
    - Is the CVD screening tool user friendly to most people and why?
    - What could be improved?
    - What would those improvements mean for the:
        a) Patients/clients
        b) Aboriginal and Torres Strait Islander Health Workers /Practitioners
        c) Clinicians

### CONSUMER TOPIC GUIDE

**Key topics (and prompts)**

- Yarning workshop feedback
    - How did you find the workshop?
    - What would have been better?
    - Did you feel involved/interested?

- Experiences with health professionals
    - Can you describe a good experience with a health professional?
    - What happened last time you saw a doctor?
    - What happened last time you saw a health worker?
    - How much input do you like to have when you talk about a health issue?

- Support for CVD prevention decisions
    - What support do you need to make decisions about heart disease prevention?
    - How do you understand your risk of heart disease?
    - What support do you have in your community to have a healthy lifestyle?
    - What information do you want about medication to prevent heart disease?

### GENERAL PRACTITIONER TOPIC GUIDE

**Key topics (and prompts)**

- GP factors
  - Do you use SDM? If so, why? Does it reflect what you were taught in medical school?
  - Do you use decision support tools in SDM? What sort of format do you find most accessible/ appropriate for patients?
  - Can you think of any decision aids for a particular medical condition that you find particularly useful?

- Patient factors
  - Are your patients responsive to SDM and its resources?
  - How do you manage patients who may feel uncertain or overwhelmed by the use of SDM resources?
  - How do you manage patients who don't want any part of SDM "you decide, *you're* the doctor"?
  - Is SDM always appropriate? Are there situations where you feel it is not appropriate?
  - Do you work with people of Aboriginal and Torres Strait Islander background?
    - How do you adapt your SDM resources to patients of Aboriginal and Torres Strait Islander background? Does this depend on other factors eg age, location, language background?
    - Is there a difference in how patients receive SDM resources in rural/remote versus urban areas?
    - Is SDM always appropriate? Are there situations where you feel it is not appropriate?

- Barriers and facilitators to SDM
  - Time constraints (incorporating SDM into standard consults)
  - Financial constraints
    - What kind of billing does your practice do?
    - Does SDM affect your billing? How?
    - Financial viability of using SDM resources for patient decision support
  - Practice structure
    - Involvement of practice nurse in SDM and CVD risk assessment/management

### THINK ALOUD INTERVIEWS WITH CONSUMERS AND GPs

What we'd like you to do is 'think aloud' about what you are doing as you look at the website, so we can understand the process you went through and what you thought about it. To help you get used to thinking aloud, we will do a practice exercise first, which is not related to heart disease.

I won't be able to answer any questions about the actual task, just do whatever you think is best, and make sure you continue to think aloud the entire time. I will prompt you to keep talking if you are silent for more than a few seconds. I will also ask you some questions at the end. Does that sound ok? Do you have any questions?
