## Supplementary file 2 for "The Heart Health Yarning Tool: co-designing a shared decision-making tool for cardiovascular disease prevention and risk management"

Supplementary file 2: Heart Health Yarning Tool website content.

Website: https://heartyarningtool.com/

Artwork: *Healthy Heart Communities* by Tyrown Waigana

### Heart Health

#### Yarning Tool

This website supports health professionals to make shared decisions about heart health checks with Aboriginal and Torres Strait Islander people. It is based on a culturally appropriate model of shared decision making that was codesigned with communities. The short video below explains key features.

It can be used:

- as a training tool for health professionals
- as a conversation guide in consultations
- to print patient summaries about heart health

STEP 1
To learn more about
how to use this tool

Learn more

STEP 2
Calculate the risk level of
the community member

Risk calculator

STEP 3
Get started

Next

This short video explains how to use the
Heart Health Yarning Tool in practice.

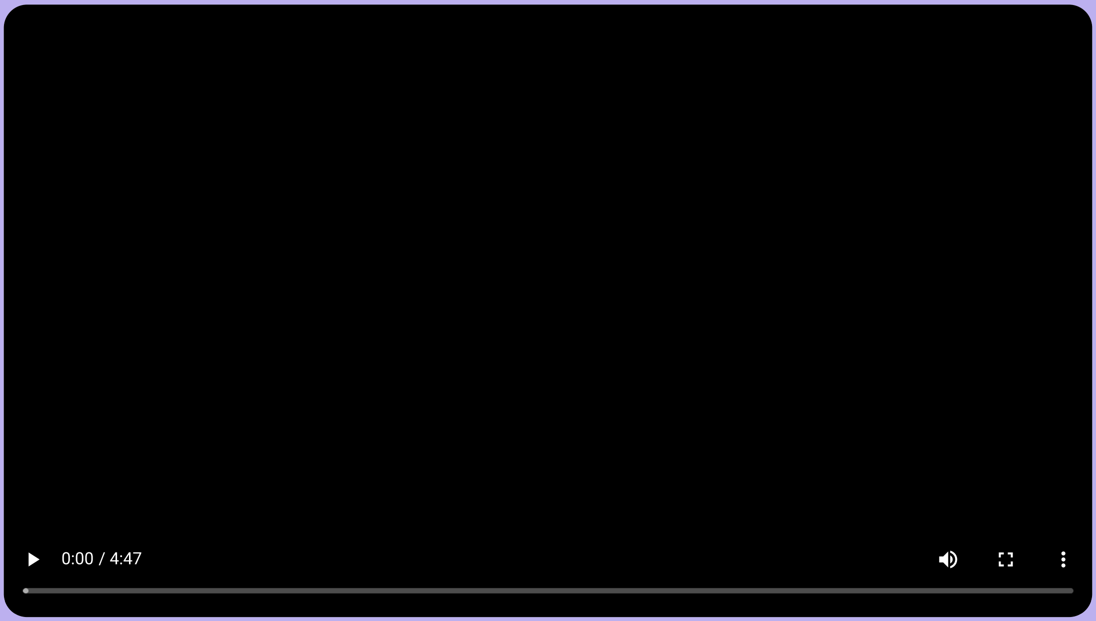


Click here to skip to high risk information:
https://heartyarningtool.com/choices-high-risk/

We acknowledge and pay our respects to Aboriginal and Torres Strait Islander Peoples as the Traditional Custodians of the Lands and Seas where we work and live.

This resource was developed by the CHAT-GP team at The University of Sydney and the Enhancing Chronic Disease Care Team at The Australian National University.
For any questions, feedback or further information on this resource, please contact

Design and website build by Saltwater People.

### How to use this tool

This website supports health professionals to make shared decisions about heart health checks with Aboriginal and Torres Strait Islander people, in a culturally appropriate way.

**Uses for health professionals:** The website can be used as a training tool for health professionals, or as a conversation guide in consultations. There are also printable* patient summaries on each page. The tool provides different information for low, medium and high risk guideline categories, and different stages of the decision making process.

**Shared decision making model:** The structure is based on the Finding Your Way model of shared decision making, which was codesigned with communities. Shared decision making means taking individual values and preferences into account, along with the evidence, to come to a shared decision. It is part of the national clinical standards.

**Heart Health Check guidelines:** The tool is designed to support Heart Health Checks based on the guidelines at https://www.cvdcheck.org.au. Once you know the level of risk, you can use this tool to guide decision making about what to do next to reduce the risk of heart disease.

* To print responses to interactive tools using a browser you will need to enable the background graphics option in your print settings. Alternatively, you can print a PDF from each webpage and fill out interactive components on the printed page.

how to use this tool

Learn more

### Welcome to the healthy heart tool

**This website is based on the Finding Your Way model of shared decision making. You will be prompted to consider cultural factors as well as discussing heart risk and prevention options.**

**Click here to skip to high risk information: https://heartyarningtool.com/choices-high-risk/**

Choose the appropriate risk category to start the conversation

Low Risk | Medium Risk | High Risk

**Click here to access the Australian CVD risk calculator**

We acknowledge and pay our respects to Aboriginal and Torres Strait Islander Peoples as the Traditional Custodians of the Lands and Seas where we work and live.

This resource was developed by the CHAT-GP team at The University of Sydney and the Enhancing Chronic Disease Care Team at The Australian National University.
For any questions, feedback or further information on this resource, please contact

Design and website build by Saltwater People.

### Welcome to the healthy heart tool

What matters to you?

Select the overarching principles to consider for shared decision making with community members.

Go    Skip

Click here to access the Australian CVD risk calculator

This website uses the Finding Your Way model of shared decision making to guide conversations about cardiovascular disease prevention. It was codesigned with input from Aboriginal And Torres Strait Islander Health Workers/Practitioners and community members.

We acknowledge and pay our respects to Aboriginal and Torres Strait Islander Peoples as the Traditional Custodians of the Lands and Seas where we work and live.

This resource was developed by the CHAT-GP team at The University of Sydney and the Enhancing Chronic Disease Care Team at The Australian National University.
For any questions, feedback or further information on this resource, please contact

Design and website build by Saltwater People.

### What matters to you?

**Here are some things you can yarn about with your health professional to help them understand what's important to you.**

**Tick/select the ones that are important to you.**

- [ ] Feeling safe
- [ ] Feeling trusted
- [ ] Culture: connecting to land and sea
- [ ] Physical and spiritual connections
- [ ] Physical Social and Emotional Wellbeing
- [ ] Sharing your stories
- [ ] Sharing your truths
- [ ] Personal experiences
- [ ] Community, family and kinship

**See your GP if you have questions or concerns about your Heart Health**

This tool uses the "Finding Your Way" model of shared decision making to guide conversations about cardiovascular disease prevention. It was codesigned with input from Aboriginal And Torres Strait Islander Health Workers/Practitioners and community members. See the "Finding Your Way" guide here: www.aci.health.nsw.gov.au/shared-decision-making

This resource was developed by the CHAT-GP team at The University of Sydney and the Enhancing Chronic Disease Care Team at The Australian National University.

For any questions, feedback or further information on this resource, please contact

### Some things to yarn about

#### Choose a place to start the conversation

- Decisions
- Options
- Support
- Family or Friends
- Ways of knowing being and doing
- Weigh up the odds
- Take a break
- Next steps

**See your GP if you have questions or concerns about your Heart Health**

This tool uses the “Finding Your Way” model of shared decision making to guide conversations about cardiovascular disease prevention. It was codesigned with input from Aboriginal And Torres Strait Islander Health Workers/Practitioners and community members. See the “Finding Your Way” guide here: www.aci.health.nsw.gov.au/shared-decision-making

This resource was developed by the CHAT-GP team at The University of Sydney and the Enhancing Chronic Disease Care Team at The Australian National University.

For any questions, feedback or further information on this resource, please contact

### Decisions

Think about the lifestyle or medication options you want to consider. Yarn with your health professional about which ones might be best for you right now.

**How are you feeling about these options?**

Quit smoking

Back Next

**What questions do you have?**

Are you ready to make a decision or do you need to take a break?

"I'd like to hear from a doctor's point of view obviously because that's their profession but I also want them to hear what I'm saying and be able to answer it in a civilian way so it's understandable…not just chucking big words around…make you understand so you don't go home with a worried mind."

– Community member

We acknowledge and pay our respects to Aboriginal and Torres Strait Islander Peoples as the Traditional Custodians of the Lands and Seas where we work and live.

This resource was developed by the CHAT-GP team at The University of Sydney and the Enhancing Chronic Disease Care Team at The Australian National University.
For any questions, feedback or further information on this resource, please contact

Design and website build by Saltwater People.

Download summary

Print page

### Decisions

Think about the lifestyle or medication options you want to consider. Yarn with your health professional about which ones might be best for you right now.

**How are you feeling about these options?**

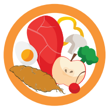

**Change diet**

Back

Next

**What questions do you have?**

Are you ready to make a decision or do you need to take a break?

**"I'd like to hear from a doctor's point of view obviously because that's their profession but I also want them to hear what I'm saying and be able to answer it in a civilian way so it's understandable…not just chucking big words around…make you understand so you don't go home with a worried mind."**

– Community member

We acknowledge and pay our respects to Aboriginal and Torres Strait Islander Peoples as the Traditional Custodians of the Lands and Seas where we work and live.

This resource was developed by the CHAT-GP team at The University of Sydney and the Enhancing Chronic Disease Care Team at The Australian National University.
For any questions, feedback or further information on this resource, please contact

Design and website build by Saltwater People.

### Decisions

Think about the lifestyle or medication options you want to consider. Yarn with your health professional about which ones might be best for you right now.

**How are you feeling about these options?**

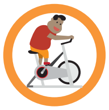

**More exercise**

Back Next

**What questions do you have?**

Are you ready to make a decision or do you need to take a break?

**"I'd like to hear from a doctor's point of view obviously because that's their profession but I also want them to hear what I'm saying and be able to answer it in a civilian way so it's understandable…not just chucking big words around…make you understand so you don't go home with a worried mind."**

– Community member

### Decisions

Think about the lifestyle or medication options you want to consider. Yarn with your health professional about which ones might be best for you right now.

**How are you feeling about these options?**

Cholesterol medication

Back
Next

**What questions do you have?**

Are you ready to make a decision or do you need to take a break?

"I'd like to hear from a doctor's point of view obviously because that's their profession but I also want them to hear what I'm saying and be able to answer it in a civilian way so it's understandable…not just chucking big words around…make you understand so you don't go home with a worried mind."

– Community member

We acknowledge and pay our respects to Aboriginal and Torres Strait Islander Peoples as the Traditional Custodians of the Lands and Seas where we work and live.

This resource was developed by the CHAT-GP team at The University of Sydney and the Enhancing Chronic Disease Care Team at The Australian National University.
For any questions, feedback or further information on this resource, please contact

Design and website build by Saltwater People.

### Decisions

Think about the lifestyle or medication options you want to consider. Yarn with your health professional about which ones might be best for you right now.

**How are you feeling about these options?**

**Blood pressure medication**

Back Next

**What questions do you have?**

Are you ready to make a decision or do you need to take a break?

**"I'd like to hear from a doctor's point of view obviously because that's their profession but I also want them to hear what I'm saying and be able to answer it in a civilian way so it's understandable…not just chucking big words around…make you understand so you don't go home with a worried mind."**

– Community member

We acknowledge and pay our respects to Aboriginal and Torres Strait Islander Peoples as the Traditional Custodians of the Lands and Seas where we work and live.

This resource was developed by the CHAT-GP team at The University of Sydney and the Enhancing Chronic Disease Care Team at The Australian National University.
For any questions, feedback or further information on this resource, please contact

Design and website build by Saltwater People.

### Options

#### 20% High risk

**Here's an example of a person with high 20% risk**
(This may be higher or lower than your risk).

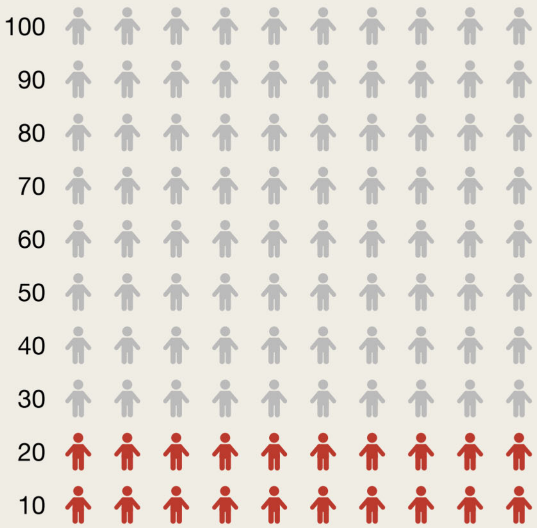


It means that out of 100 people with the same risk factors, 20 of those people will have a heart attack or stroke in the next 5 years if they don't take action.

**See your GP if you have questions or concerns about your Heart Health**

### Options

#### Take cholesterol medication

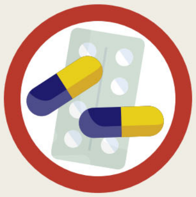

#### 15% High risk

If you reduce your cholesterol, you can reduce your risk of heart problems. This person reduced their LDL or "bad" cholesterol by 1mmol/L. Their risk changed from 20% to 15%. This means they have a better chance of keeping their heart healthy.

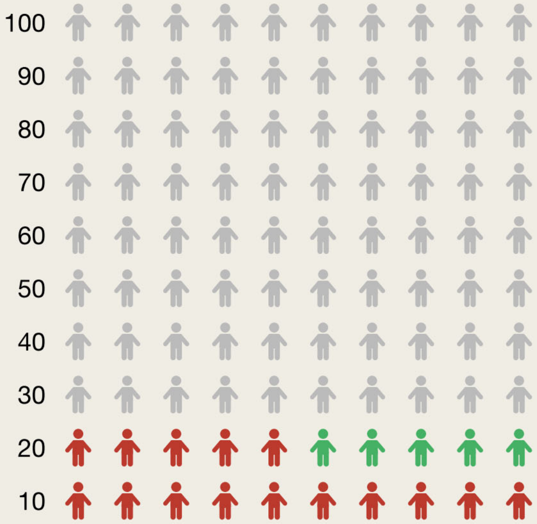


It means that out of 100 people with the same risk factors, 15 of those people will have a heart attack or stroke in the next 5 years if they don't take action.

**See your GP if you have questions or concerns about your Heart Health**

### Options

#### Take blood pressure medication and exercise

##### 16% High risk

If you reduce your blood pressure, you can reduce your risk of heart problems. This person reduced their blood pressure by 10mmHg. Their risk changed from 20% to 16%. This means they have a better chance of keeping their heart healthy.

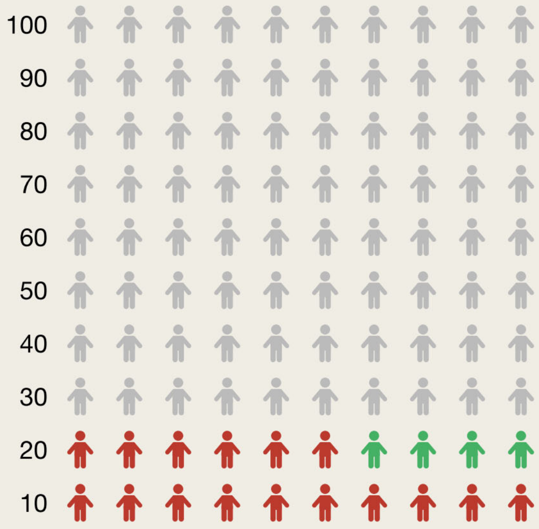


It means that out of 100 people with the same risk factors, 16 of those people will have a heart attack or stroke in the next 5 years if they don't take action.

**See your GP if you have questions or concerns about your Heart Health**

### Options

#### Quit smoking

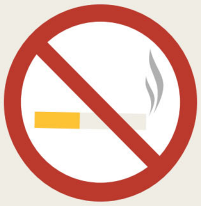

#### 12% High risk

**If you quit smoking, you can reduce your risk of heart problems.**
This person quit smoking. Their risk changed from 20% to 12%.
This means they have a better chance of keeping their heart healthy.

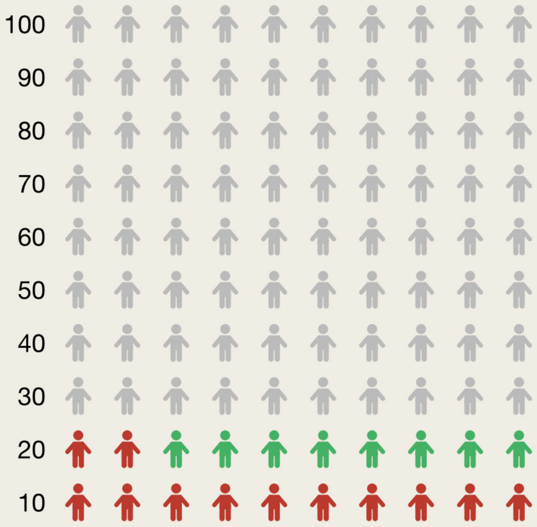


It means that out of 100 people with the same risk factors, 12 of those people will have a heart attack or stroke in the next 5 years if they don't take action.

**See your GP if you have questions or concerns about your Heart Health**

### Support

You have more in your life than just your heart health needs, talk about what else is important to your social and emotional wellbeing.

You can yarn to family, friends, health professionals and other people in your community about what you might need. Support can come from a team of health professionals, not just your health worker or doctor.

It's important for the people who are looking after you to know what works well for you, to support you in the best way for you.

**"We used to go to walking groups, and they said you might be interested in endocrinologist via telehealth, and it's been beneficial. It's something that I would not have considered, and I found it a very beneficial referral."**

**- Community member**

**See your GP if you have questions or concerns about your Heart Health**

This tool uses the "Finding Your Way" model of shared decision making to guide conversations about cardiovascular disease prevention. It was codesigned with input from Aboriginal And Torres Strait Islander Health Workers/Practitioners and community members. See the "Finding Your Way" guide here: www.aci.health.nsw.gov.au/shared-decision-making

This resource was developed by the CHAT-GP team at The University of Sydney and the Enhancing Chronic Disease Care Team at The Australian National University.

For any questions, feedback or further information on this resource, please contact

### Family and Friends

Talking about your family and friends, your country and their experiences of heart and vascular issues can be helpful.

Yarn with your family and friends about your options. It is okay to make these decisions together with family.

Making decision with others is ok. You can bring family or friends to the doctor to help you make decisions.

You might all make some changes together to improve your health e.g., you might all go for a walk together every day.

Your family may have had to find solutions to improve their health in the past which may work for you too. They may have some tips they can share with you.

**See your GP if you have questions or concerns about your Heart Health**

This tool uses the "Finding Your Way" model of shared decision making to guide conversations about cardiovascular disease prevention. It was codesigned with input from Aboriginal And Torres Strait Islander Health Workers/Practitioners and community members. See the "Finding Your Way" guide here: www.aci.health.nsw.gov.au/shared-decision-making

This resource was developed by the CHAT-GP team at The University of Sydney and the Enhancing Chronic Disease Care Team at The Australian National University.

For any questions, feedback or further information on this resource, please contact

### Ways of knowing, being and doing

You are the expert about your story and your body. Nobody knows your journey better than you do, and everyone's journey is different. Here are some questions you can yarn about with your health professional to help them understand what's important to you.

- Where do you feel safe to talk about your heart health?
- Who do you trust to talk about your heart health with?
- Are there any stories you want to share about your heart health?
- What is true for you about your heart health?
- How can your community and family support your heart health?
- How can physical and spiritual connections help you on your heart journey?
- How can your culture and connection to land and sea help you on your heart journey?
- How might your personal experiences affect your heart health?
- What do you need to support your physical, social and emotional wellbeing?

**"I guess you know being Indigenous…it's hard for us to talk to non-Indigenous people…it's good to talk with mob you know who could break it down for us and explain it. You felt comfortable enough to ask more questions then."**

**- Community member**

**See your GP if you have questions or concerns about your Heart Health**

This tool uses the "Finding Your Way" model of shared decision making to guide conversations about cardiovascular disease prevention. It was codesigned with input from Aboriginal And Torres Strait Islander Health Workers/Practitioners and community members. See the "Finding Your Way" guide here: www.aci.health.nsw.gov.au/shared-decision-making

This resource was developed by the CHAT-GP team at The University of Sydney and the Enhancing Chronic Disease Care Team at The Australian National University.

For any questions, feedback or further information on this resource, please contact

### Weigh up the odds

There are different ways that you can improve your heart health. There will be good and bad things about each of these options.

Think about which heart health options are best for you, your personal circumstances and what support you have from family, friends and your community.

**What about these options might work for you?**

Talk about these options with your health professional.

Lifestyle change options

Medication options

**"She told me the pros and cons of it and the cost of it because it wasn't covered ...the effects of it. I didn't do much research into it and then when she explained it to me from A to B, then it kind of made sense"**

– Community member

We acknowledge and pay our respects to Aboriginal and Torres Strait Islander Peoples as the Traditional Custodians of the Lands and Seas where we work and live.

This resource was developed by the CHAT-GP team at The University of Sydney and the Enhancing Chronic Disease Care Team at The Australian National University.
For any questions, feedback or further information on this resource, please contact

Design and website build by Saltwater People.

### Lifestyle change options

If your health professional said you could change your lifestyle to improve your heart health, pick one thing to focus on for now and ask them what extra support you can get in your community:

**Smoke less**

- ☐ Call Aboriginal Quitline
- ☐ Use nicotine gum or patches
- ☐ Reduce number of cigarettes
- ☐ Other

**Diet changes**

- ☐ Have less sugary drinks
- ☐ Eat more healthy snacks
- ☐ Use less salt in cooking
- ☐ Other

**Be more active**

- ☐ Walk more
- ☐ Exercise at home
- ☐ Join a local sport team
- ☐ Other

**Do you want to see how much quitting smoking can reduce your chance of a heart attack or stroke?**

### Quit smoking

#### 12% High risk

**If you quit smoking, you can reduce your risk of heart problems.**

This person quit smoking. Their risk changed from 20% to 12%. This means they have a better chance of keeping their heart healthy.

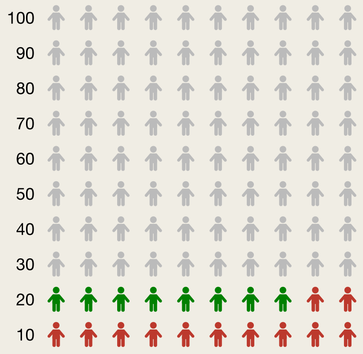


**Do you want to make a plan for how you can change your habits?**

Yes

We acknowledge and pay our respects to Aboriginal and Torres Strait Islander Peoples as the Traditional Custodians of the Lands and Seas where we work and live.

This resource was developed by the CHAT-GP team at The University of Sydney and the Enhancing Chronic Disease Care Team at The Australian National University.
For any questions, feedback or further information on this resource, please contact

Design and website build by Saltwater People.

Download summary

### Change your habits

**Planning tool**

We can start by changing one habit at a time.

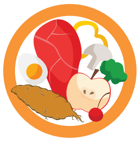

##### Change your snacking habits

Go

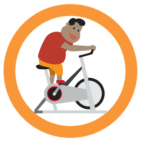

##### Change your exercise habits

Go

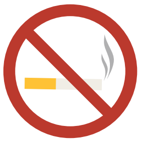

##### Change your smoking habits

Go

We acknowledge and pay our respects to Aboriginal and Torres Strait Islander Peoples as the Traditional Custodians of the Lands and Seas where we work and live.

This resource was developed by the CHAT-GP team at The University of Sydney and the Enhancing Chronic Disease Care Team at The Australian National University.
For any questions, feedback or further information on this resource, please contact

Design and website build by Saltwater People.

### Change your habits

#### Snacking habits

##### I often eat unhealthy snacks when...

- ☐ I am busy or stressed
- ☐ I am sad
- ☐ I am happy
- ☐ I have a craving
- ☐ The snack is right in front of me
- ☐ I have arrived home
- ☐ I am bored
- ☐ I start with one piece but then keep eating
- ☐ I want to reward myself
- ☐ Someone offers me a snack
- ☐ People around me are eating
- ☐ I am drinking coffee or tea
- ☐ I am in front of a TV or computer
- ☐ I am drinking alcohol
- ☐ It is part of a celebration or special event

#### Possible solutions

- ☐ Do a chore or a task
- ☐ Eat a piece of fruit
- ☐ Listen to my favourite music or podcast
- ☐ Eat a smaller amount
- ☐ Chat to someone for five minutes
- ☐ Drink a large glass of water
- ☐ Eat fresh vegetables an dip
- ☐ Move the snack into the cupboard
- ☐ Drink tea

### Change your habits

#### Exercise habits

##### I often don’t want to exercise when...

- [ ] The weather is bad
- [ ] I am too tired
- [ ] I don’t feel I have the time to
- [ ] I am in pain
- [ ] I feel it will be boring
- [ ] I don’t feel motivated
- [ ] I have to go on my own
- [ ] I can’t see the physical benefits from exercising
- [ ] I am too busy
- [ ] I feel lazy
- [ ] I don’t have access to equipment or facilities
- [ ] I have work to do
- [ ] Family events of situations get in the way
- [ ] I feel out of shape

#### Possible solutions

- [ ] Ask someone to come with me
- [ ] Remind myself of the benefits of exercising
- [ ] Try a new way of exercising
- [ ] Think of how my inactivity affects those around me
- [ ] Do a short exercise at home
- [ ] Tell myself that exercising will make me a healthier person
- [ ] Remind myself that I will feel better afterwards
- [ ] Just start with 10 minutes of exercise
- [ ] Set a timer for 5 minutes and then exercise

### Change your habits

#### Smoking habits

##### I often smoke when...

- [ ] I am driving
- [ ] I am sad
- [ ] Someone offers me a cigarette
- [ ] I am frustrated
- [ ] I am stressed
- [ ] I have just eaten a meal
- [ ] I am working or studying
- [ ] Someone is smoking in front of me
- [ ] I am happy
- [ ] I have withdrawal symptoms
- [ ] I am drinking coffee or tea
- [ ] A negative event happens in my area
- [ ] It's the morning
- [ ] I am drinking alcohol

#### Possible solutions

- [ ] Chew some gum
- [ ] Listen to my favourite music or a podcast
- [ ] Drink a large glass of water
- [ ] Remember how good I'll feel about myself if I don't smoke
- [ ] Chat to someone for 5 minutes
- [ ] Seek out someone who listens when I need to talk about my smoking
- [ ] Think of what this means for people I care about
- [ ] Eat a healthy snack
- [ ] Remember the benefits of stopping
- [ ] Do some exercises like 10 star jumps
- [ ] Use my hands for something else (like knitting or playing a video game)
- [ ] Use mouthwash

### Lifestyle change options

If your health professional said you could change your lifestyle to improve your heart health, pick one thing to focus on for now and ask them what extra support you can get in your community:

**Smoke Less**

- [ ] Call Aboriginal Quitline
- [ ] Use nicotine gum or patches
- [ ] Reduce number of cigarettes
- [ ] Other

**Diet changes**

- [ ] Have less sugary drinks
- [ ] Eat more health snacks
- [ ] Use less salt in cooking
- [ ] Other

**Be more active**

- [ ] Walk more
- [ ] Exercise at home
- [ ] Join a local sport team
- [ ] Other

**Do you want to make a plan for how you can change your habits?**

**See your GP if you have questions or concerns about your Heart Health**

This tool uses the "Finding Your Way" model of shared decision making to guide conversations about cardiovascular disease prevention. It was codesigned with input from Aboriginal And Torres Strait Islander Health Workers/Practitioners and community members. See the "Finding Your Way" guide here: www.aci.health.nsw.gov.au/shared-decision-making

This resource was developed by the CHAT-GP team at The University of Sydney and the Enhancing Chronic Disease Care Team at The Australian National University.

For any questions, feedback or further information on this resource, please contact

Download summary

Print page

### Medication options

If your health professional said medication could improve your heart health, pick one thing to focus on for now and ask them what extra support you can get in your community.

| Take cholesterol medication<br>*Take medicine every day to reduce your cholesterol.* | Blood pressure medication<br>*Take medicine every day to reduce your blood pressure.* |
|---|---|
| **Pros:** Reduces your chance of a heart attack or stroke. | **Pros:** Reduces your chance of a heart attack or stroke. |
| **Cons:** You may get side effects like muscle aches. | **Cons:** You may get side effects like feeling dizzy or fainting. |
| Is this something you might consider?<br>Yes/no | Is this something you might consider?<br>Yes/no |

**Do you want to see how much medication can reduce your chance of a heart attack or stroke?**

#### Take cholesterol medication

**15% High risk**

If you take a cholesterol lowering medication you could gradually reduce your absolute risk from 20% to 15%. Your new risk is considered high.

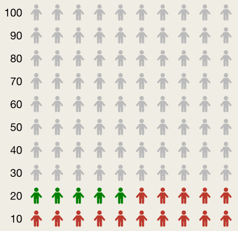


It means that out of 100 people with the same risk factors, 15 of those people will have a heart attack or stroke in the next 5 years if they don't take action.

**Do you want more information** Yes

We acknowledge and pay our respects to Aboriginal and Torres Strait Islander Peoples as the Traditional Custodians of the Lands and Seas where we work and live.


This resource was developed by the CHAT-GP team at The University of Sydney and the Enhancing Chronic Disease Care Team at The Australian National University.
For any questions, feedback or further information on this resource, please contact

Design and website build by Saltwater People.

### Blood pressure medication

#### More information

**Blood pressure medication decision aid**

Australian guidelines recommend blood pressure-lowing medication for those at high risk. It may also be recommended for moderate risk if you have extra risk factors or if lifestyle change is not effective after 3-6 months. This involves taking a pill every day for as long as you and your doctor agree that the benefits outweigh the harms.

The main medication used are:

- ACE inhibitor
- Angiotensin Receptor Blocker
- Calcium Channel Blocker
- Low dose thiazide or thiazide-like diuretic

| | Benefits | Harm |
|---|---|---|
| **Take action:**<br><br>**Start blood pressure medication** | 79 in 1000 people have a CVD event while taking a blood pressure lowering medication compared to 82 in 1000 taking a placebo pill. | Hypotension (low blood), 14 in 1000 aiming for <140mm Hg may to 23 in 1000 aiming for <120mm Hg.<br>Syncope (Fainting) 17 in 1000<br>Aiminypg for <120mm Hg.<br>Acute kidney injury / renal failure 25 in 1000 aiming for <140mm Hg may present with hypotension |
| **Do nothing:**<br><br>**Don't take medication** | You will not be at risk of the side effects of taking medication. | Your CVD risk will continue to increase as your age if not other actions are taken to reduce your blood pressure and cholesterol. |

**Based on the information I am:**

Leaning towards medication — Leaning against medication

### Cholesterol medication

#### More information

##### Cholesterol medication decision aid

Australian guidelines recommend cholesterol lowering medication for those at high risk. It may also be recommended for moderate risk if you have extra risk factors or if lifestyle change is not effective after 3-6 months. This involves taking a pill every day for as long as you and your doctor agree that the benefits outweigh the harms. The dose and type of medication may need to be adjusted depending how well you tolerate the initial pill, and what effects this has on your LDL (low density lipoprotein) cholesterol the “bad” cholesterol. The most common type is called Statin.

| | Benefits | Harm |
|---|---|---|
| **Take action:**<br><br>**Start choleserol medication** | 92 in 1000 people have a CVD event while taking a placebo pill. 17 in 1000 people have a stroke event while taking statin compared to 22 in 1000 taking a placebo pill. | Cost: Recurring cost for daily pill.<br><br>Side effects: Commonly reported events include muscle aches, arthritis, nausea, constipation, diarrhoea, elevated liver enzymes, kidney disorders and rhabdomyolysis (serious muscle injury), but the rates are about the same for statins vs placebo. 24 in 1000 people may develop diabetes when on statin compared to 28 in 1000 on a placebo pill. You may also need to have your liver monitored (via a blood sample) for a couple of months when you first start statin |
| **Do nothing:**<br><br>**Don't take medication** | You will not be at risk of the side effects of taking medication. | Your CVD risk will continue to increase as your age if not other actions are taken to reduce your blood pressure and cholesterol. |

**Based on the information I am:**

Leaning towards medication — Leaning against medication

### Take a break

You might need more time to think about the options, or yarn with family and friends. It's ok to come back another time if you're not ready to make a decision yet.

If you don't feel like you're in a safe place to yarn with someone you trust, you can talk to a different health professional. You can ask about seeing an Aboriginal and Torres Strait Islander Health Worker or Practitioner if you don't feel comfortable with your GP or nurse. You can ask to see a male or female health professional to help you feel more comfortable.

**"You're not going to get all this done in one session. So, being able to have that follow-up, being having that ongoing care, having the ongoing talking about, sharing our stories - having something you can take away and being followed up is very important."**

**- Community member**

**See your GP if you have questions or concerns about your Heart Health**

This tool uses the "Finding Your Way" model of shared decision making to guide conversations about cardiovascular disease prevention. It was codesigned with input from Aboriginal And Torres Strait Islander Health Workers/Practitioners and community members. See the "Finding Your Way" guide here: www.aci.health.nsw.gov.au/shared-decision-making

This resource was developed by the CHAT-GP team at The University of Sydney and the Enhancing Chronic Disease Care Team at The Australian National University.

For any questions, feedback or further information on this resource, please contact

### Next steps

When you are trying to make changes for your health some things work well and some things take more time to put into place.

Some things you try may not work at all for you, at first – talk to your healthcare team and family about how they can support you to help you find what is best for you.

Checking in is important and bring the people who support you on your health journey.

**"This is what stuck with me, that he was naturally curious, and he actually asked me, "What are the challenges, to you, getting a screen right here, right now?" Instead of giving me a lecture what he was saying was, "What can I do to help you to consider a screen?"**

**- Community member**

**See your GP if you have questions or concerns about your Heart Health**

This tool uses the "Finding Your Way" model of shared decision making to guide conversations about cardiovascular disease prevention. It was codesigned with input from Aboriginal And Torres Strait Islander Health Workers/Practitioners and community members. See the "Finding Your Way" guide here: www.aci.health.nsw.gov.au/shared-decision-making

This resource was developed by the CHAT-GP team at The University of Sydney and the Enhancing Chronic Disease Care Team at The Australian National University.

For any questions, feedback or further information on this resource, please contact

### Risk check

#### Cardiovascular disease (CVD)

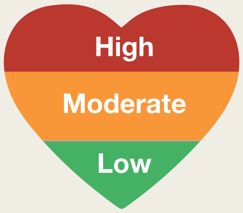


##### Your CVD risk is ________________

This means that if there were 100 people like you we would expect _________ of them to have a heart attack or stroke within the next 5 years.

##### You can reduce your risk of a heart attack or stroke

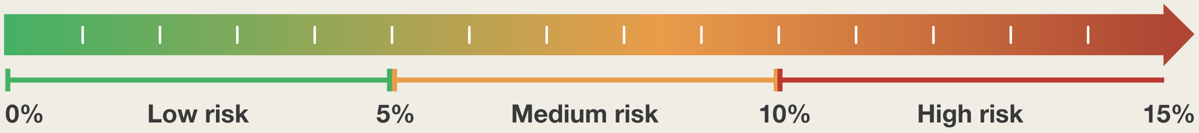


##### 8 things that can contribute to your current CVD risk

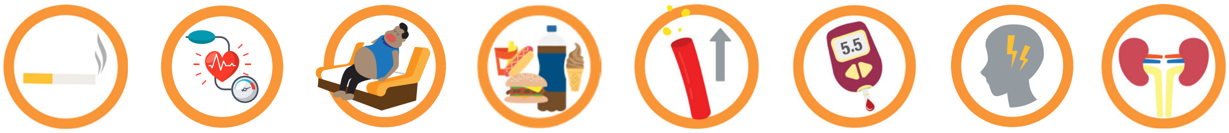


- Smoking
- High blood pressure
- Being inactive
- Unhealthy diet
- High cholesterol
- Diabetes
- Depression & social isolation
- Kidney disease

##### To improve your heart health

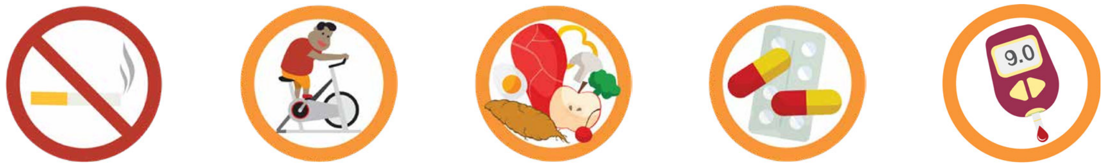


- Quit Smoking
- Do more physical activity
- Eat a healthy, balanced diet
- Take blood pressure & cholesterol lowering medication
- Manage your diabetes

**Let's talk about services you can access to help you reduce your risk of heart attack or stroke, and how to have a healthier lifestyle!**

### Yarn about your heart health

#### Here are some things you can yarn about with a trusted health professional.

**Lifestyle changes and medicine can help your heart.**

**Ask questions about your options to help you make a decision.**

**Yarn about what else is important to your social and emotional wellbeing.**

**Yarn about the next steps you can take.**

**It's ok to come back another time if you're not ready to make a decision.**

**Yarn about the pros and cons of the options.**

**Share stories about your country, your family and their heart journeys.**

**Share your knowledge about what's important for you.**

#### What matters to you?

**Here are some questions you can yarn about with a trusted health professional. This will help them understand what's important to you.**

- [ ] Where do you feel safe to talk about your heart health?
- [ ] Who do you trust to talk about your heart health with?
- [ ] Are there any stories you want to share about your heart health?
- [ ] What is true for you about your heart health?
- [ ] How can your community and family support your heart health?
- [ ] How can physical and spiritual connections help you on your heart journey?
- [ ] How can your culture and connection to land and sea help you on your heart journey?
- [ ] How might your personal experiences affect your heart health?
- [ ] What do you need to support your physical, social and emotional wellbeing?
